# Childhood Trauma Exposure Increases Long COVID Risk

**DOI:** 10.1101/2022.02.18.22271191

**Authors:** Alicia W. Villanueva Van Den Hurk, Cady Ujvari, Noah Greenspan, Dolores Malaspina, Xavier F. Jimenez, Julie Walsh-Messinger

## Abstract

**Background:** A proportion of those who contract COVID-19 will develop long COVID (i.e., symptoms that persist for three months or more). Childhood trauma contributes to a pro-inflammatory state in adulthood evidenced by high morbidity and early mortality, but it has not yet been investigated as a risk factor for long COVID.

**Methods:** Participants (N=338) completed online measures of premorbid health, COVID-19 positivity, symptoms, recovery, depression, anxiety, and post-traumatic stress disorder (PTSD). Questionnaires about childhood and recent traumatic experiences were completed by half of the sample (N=162).

**Results:** Fifty-three percent of participants developed long COVID, of whom over 60% endorsed exercise intolerance and protracted myalgias, headaches, brain fog, and shortness of breath. Participants who experienced at least one childhood traumatic event were 3-fold more likely to develop the syndrome (OR=3.11, 95% CI, 1.49 to 6.48), while risk was nearly 6-fold increased for two or more events (OR=5.67, CI, 2.44 to 13.13). Regression models showed childhood trauma (OR=5.32, CI, 1.44 to 19.68), older age (OR=1.11, CI, 1.06 to 1.16), female sex (OR=4.02, CI, 1.34 to 12.12), along with chest pain (OR=8.77, CI, 2.80 to 27.43), brain fog (OR=3.33, CI, 1.16 to 9.57) and phantosmia (OR=5.90, CI, 1.40 to 24.86) during acute illness accurately classified long COVID status in 87% of participants.

**Interpretations:** Early adversity is a risk-factor for long COVID, likely due to altered immune response, central sensitization, and peripheral dysfunction. Childhood trauma, a crucial social determinant of health, should be routinely assessed in COVID-19 survivors and may aid in determining prognosis.

## INTRODUCTION

The severe acute respiratory syndrome coronavirus 2 (SARS-COV-2), which causes the COVID-19 disease, has infected over 412 billion people worldwide since it was first identified in Wuhan in December 2019.^1^ Although the long-term physical and psychological effects of COVID-19 remain unknown, it is now clear that a proportion of survivors experience protracted respiratory, cardiovascular, neurologic, dermatologic, and/or gastrointestinal symptoms and complications following acute illness.^2^ The syndrome is highly variable in presentation, although fatigue, dyspnea, cough, chest pain, headache, chemosensory impairment, diarrhea, and muscle pain are reported most frequently.

Universal terminology for this post-viral syndrome is not yet recognized, but it is frequently referred to as ‘post-acute sequalae of SARS-CoV-2 (PASC),’ ‘long-haul COVID,’ and ‘long COVID.’ There is also a lack of consensus on timeframe. The United States Center for Disease Control (CDC)^3^ recognizes long COVID after persistent symptoms at least four weeks, whereas the World Health Organization (WHO)^4^ and the United Kingdom’s National Health Services (NHS)^5^ require symptoms to persist for three months or more. Herein we use the latter, more conservative definition.

It is estimated that approximately 30% to 50% of non-hospitalized COVID patients will develop the syndrome,^6-10^ with even higher rates reported in hospitalized cases.^11,12^ However, a study that employed a daily COVID-19 symptom tracking app found only two percent of participants reported symptoms three months after illness onset,^13^ suggesting rates may vary by sample ascertainment. Female sex, older age, higher body mass index (BMI), greater number of symptoms during acute illness, chemosensory impairment, and diarrhea are identified as predictors of long COVID.^6-9,12-15^

Several mechanisms may ultimately be implicated in long COVID, but excess proinflammatory cytokines (e.g., interleukin-6 (IL-6), tumor necrosis factor alpha (TNFα), c-reactive protein), interacting with chronic neuroinflammation is one proposed mechanism.^16^Increased levels of proinflammatory cytokines are linked to depression, anxiety, and post-traumatic stress disorder (PTSD),^17,18^ which may be mediated by the effect of childhood trauma and early adversity on brain development and immune response.^19^

While adverse childhood experiences are robustly associated with negative health outcomes in adulthood and premature mortality,^20^ early trauma has not yet been examined as a risk factor for long COVID. The present study investigated whether childhood trauma exposure is a risk factor for long COVID and characterized the physical and mental health consequences of the syndrome. We hypothesized that childhood trauma would predict long COVID risk, and that depression, anxiety, and PTSD severity would be higher in long COVID participants compared to those who fully recovered.

## METHODS

### Participants

Participants (N=455) were recruited via social media postings on Facebook, Twitter, and Instagram (N=150), Prolific.co (N=289), an online research participant recruitment site that prescreens and matches registrants to research studies, and a long COVID clinical treatment trial (N=16). Eligibility was determined by 1) age ≥ 18 years, 2) ability to complete questionnaires in English, 3) a positive PCR test, clinician diagnosis, or belief of SARS-CoV-2 infection based on symptoms.

Excluded from the aforementioned sample were one participant with dementia (rendering self-report unreliable), 13 who had remained symptomatic after 30 days but completed the measures < 90 days after illness onset, 28 who completed the measures <30 days post illness onset, and 75 who never received a positive PCR test or a clinician diagnosis, and did not report chemosensory impairment during acute illness, which is the strongest indictor of COVID-19 positivity.^21^

The final sample (N=338) included 158 participants (47%) who fully recovered within 30 days and 180 who developed long COVID (53%), defined as protracted symptoms and post-viral complications lasting ≥ three months after acute illness. All participants contracted COVID-19 between January 2020 and January 2021.

### Procedure

Measures for this cross-sectional study were administered online through Qualtrics (QualtricsXM, Provo, UT). A subset (N=162) completed online questionnaires about childhood and recent traumatic events. All studies were approved by the University of Dayton Institutional Review Board (IRB), and participants provided voluntary informed consent obtained in accordance with the Declaration of Helsinki.

### Measures

#### COVID-19 Questionnaire

Our COVID-19 questionnaire asked about medical and psychiatric history, COVID-19 positivity, acute symptoms, course of illness, treatment, and post-COVID symptoms and complications. Acute illness severity was assessed with questions 2a, 2b, and 2c from the *COVID Experiences (COVEX) Symptoms and Diagnoses* module.^15^

Premorbid health risk factors included cardiac and pulmonary conditions, obesity, liver and kidney disease, cancer, immunocompromised state, neurodegenerative disease, active pregnancy, current or past history of smoking, substance use disorder per CDC guidelines.^22^

#### Mental Health Measures

Depression severity was assessed with the *Patient Health Questionnaire-9* (PHQ-9)^23^ (N=322) or the *Center for Epidemiology Studies Depression Scale – Revised* (CESD-R)^24^ (N=16). Major depression was diagnosed using diagnostic criteria outlined by each measure. To allow for comparison across the two measures we converted the PHQ9 and CESD-R scores to t-scores based on the Patient-Reported Outcomes Measurement Information System (PROMIS) conversion chart.^25^ Anxiety was assessed with the *Generalized Anxiety Disorder-2 Item* (GAD-2)^26^ and the *Posttraumatic Stress Disorder (PTSD) Checklist for DSM-5* (PCL-5)^27^ assessed PTSD symptom severity. Criteria outlined by measure authors were used for PTSD and GAD diagnosis.

### Childhood and Recent Trauma

The *Childhood Traumatic Events Scale*^28^ assessed six potential traumatic experiences prior to age 17, including death of a loved one, parental divorce, physical violence exposure, sexual abuse, serious illness or injury, or other (participant specified) traumatic event. The *Recent Traumatic Events Scale*^28^ assessed the death of a loved one, relationship stressors, non-sexual violence, sexual assault, serious illness or injury, vocational stress, or other (participant specified) traumatic event in the three years prior to contracting COVID-19. For each event endorsed, participants indicated the age at which the event first occurred, and event trauma on a scale of 0 (not experienced) to 7 (extremely). Scores were then computed for childhood and recent trauma burden, by summing the event trauma ratings.

### Data Analysis

Data analysis was performed with SPSS (version 27.0). Descriptive statistics were first used to identify any features of the data (e.g., non-normal distribution, outliers, skewness) that might influence inferential methods and all variables were determined to be normally distributed. Independent samples t-tests, chi-square, Fisher’s Exact test, and Cramer’s V tested group differences in demographic and health variables, as appropriate. Pearson correlations tested associations between continuous and variables and Spearman’s rho (r_s_) was employed when one or more variables were ordinal.

Two (recovered, long COVID) x two (male, female) ANCOVAs were used to examine group differences in PTSD, depression, and generalized anxiety severity, and childhood and recent trauma burden when controlling for age. We then employed stepwise binary logistic regression with backward Wald entry to examine predictors of long COVID. Included in the first model were age, sex, number of premorbid risk factors, and acute illness symptoms. In our second model, tested only in the subsample who completed the trauma measures, we entered the significant predictors from model one along with number of childhood and recent trauma exposures.

#### Sample Size Estimation

We estimated the minimum sample size using G-Power version 3.1.9.7 and determined that samples of N=128 and N=351 and would provide 80% power to detect moderate and small effects, respectively.

## RESULTS

### Sample Characteristics and Premorbid Health

Compared to recovered participants, the long COVID group was older and predominantly female, but did not differ in race or ethnicity (Table 1). The majority of participants reported “good” (51.5%) or “excellent” (37.6%) premorbid health, with no differences between the groups (Cramer’s V=.087, p=.460). More than half of the sample (56.7%) had a least one premorbid health risk factor, with similar rates across both groups (X^2^=0.43, p=.836). Current smokers were more likely to fully recover (X^2^=4.85, p=.028), while those with obesity were more likely to develop long COVID (X^2^=10.44, p=.001).

**Table 1.**
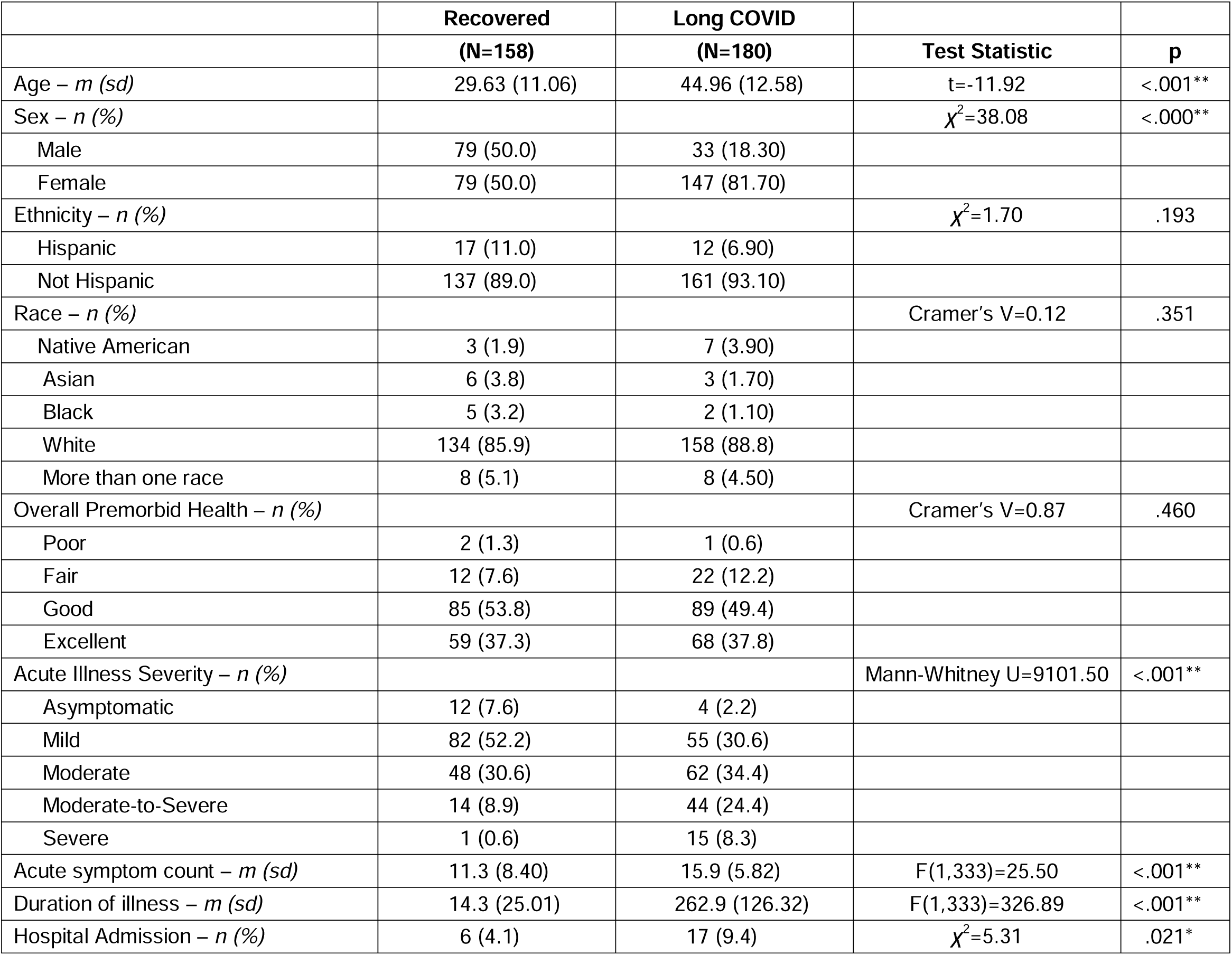
Demographic and acute illness characteristics stratified by COVID-19 recovery status.

|  | **Recovered**<br>**(N=158)** | **Long COVID**<br>**(N=180)** | **Test Statistic** | **p** |
| --- | --- | --- | --- | --- |
| Age – *m (sd)* | 29.63 (11.06) | 44.96 (12.58) | $t=-11.92$ | <.001** |
| Sex – *n (%)* | | | $\chi^2=38.08$ | <.000** |
| Male | 79 (50.0) | 33 (18.30) |  |  |
| Female | 79 (50.0) | 147 (81.70) |  |  |
| Ethnicity – *n (%)* | | | $\chi^2=1.70$ | .193 |
| Hispanic | 17 (11.0) | 12 (6.90) |  |  |
| Not Hispanic | 137 (89.0) | 161 (93.10) |  |  |
| Race – *n (%)* |  |  | Cramer's V=0.12 | .351 |
| Native American | 3 (1.9) | 7 (3.90) |  |  |
| Asian | 6 (3.8) | 3 (1.70) |  |  |
| Black | 5 (3.2) | 2 (1.10) |  |  |
| White | 134 (85.9) | 158 (88.8) |  |  |
| More than one race | 8 (5.1) | 8 (4.50) |  |  |
| Overall Premorbid Health – *n (%)* |  |  | Cramer's V=0.87 | .460 |
| Poor | 2 (1.3) | 1 (0.6) |  |  |
| Fair | 12 (7.6) | 22 (12.2) |  |  |
| Good | 85 (53.8) | 89 (49.4) |  |  |
| Excellent | 59 (37.3) | 68 (37.8) |  |  |
| Acute Illness Severity – *n (%)* |  |  | Mann-Whitney U=9101.50 | <.001** |
| Asymptomatic | 12 (7.6) | 4 (2.2) |  |  |
| Mild | 82 (52.2) | 55 (30.6) |  |  |
| Moderate | 48 (30.6) | 62 (34.4) |  |  |
| Moderate-to-Severe | 14 (8.9) | 44 (24.4) |  |  |
| Severe | 1 (0.6) | 15 (8.3) |  |  |
| Acute symptom count – *m (sd)* | 11.3 (8.40) | 15.9 (5.82) | $F(1,333)=25.50$ | <.001** |
| Duration of illness – *m (sd)* | 14.3 (25.01) | 262.9 (126.32) | $F(1,333)=326.89$ | <.001** |
| Hospital Admission – *n (%)* | 6 (4.1) | 17 (9.4) | $\chi^2=5.31$ | .021* |

Long COVID participants had more premorbid hypertension, hyperlipidemia, migraine headaches, osteoarthritis, thyroid disorder, and fibromyalgia, and tended to have higher rates of food allergies and Ehlers-Danlos syndrome (Table 2). Rates of psychiatric diagnosis did not differ between the groups, with the exception of bipolar II disorder, which, although rare, was more frequent in recovered participants.

**Table 2.** Comparison of pre and post COVID health and mental health diagnoses.

|  | Premorbid |  |  | Post-COVID |  |  |
| --- | --- | --- | --- | --- | --- | --- |
| | Recovered<br>(N=158) | Long COVID<br>(N=180) | $\chi^2$ | Recovered<br>(N=158) | Long COVID<br>(N=180) | $\chi^2$ |
| Diagnosis – *n* (%) |  |  |  |  |  |  |
| Obesity | 14 (8.9) | 39 (21.7) | 10.44** | 14 (8.9) | 41 (22.8) | 11.47** |
| Hyperlipidemia | 1 (0.6) | 15 (8.3) | 11.06** | 2 (1.3) | 24 (13.3) | 16.90** |
| Hypertension | 11 (7.0) | 28 (15.6) | <sup>F</sup>6.09* | 13 (8.4) | 33 (18.3) | 6.09* |
| Hypotension | 1 (0.6) | 5 (2.8) | <sup>F</sup>0.20 | 2 (1.3) | 11 (6.1) | 5.20 |
| Asthma | 27 (17.1) | 31 (17.2) | 0.00 | 27 (17.1) | 43 (32.1) | 2.37 |
| Cystic fibrosis | 8 (5.1) | 6 (3.3) | 0.63 | 8 (5.1) | 9 (5.0) | 0.00 |
| Thyroid disorder | 5 (3.2) | 17 (9.4) | 5.45* | 6 (3.9) | 18 (10.0) | 4.70* |
| Gastroesophageal reflux disease | 0 (0.0) | 4 (2.2) | <sup>F</sup>0.13 | 0 (0.0) | 6 (3.3) | <sup>F</sup>0.03* |
| Osteoarthritis | 2 (1.3) | 15 (8.3) | 8.80** | 2 (1.3) | 15 (8.3) | 8.80** |
| Ehlers-Danlos syndrome | 0 (0.0) | 5 (2.8) | <sup>F</sup>0.06 | 0 (0.0) | 6 (3.3) | <sup>F</sup>0.03* |
| Herpes simplex virus I | 7 (4.4) | 14 (7.8) | 1.60 | 7 (4.5) | 15 (8.3) | 1.98 |
| Eczema | 17 (10.8) | 26 (14.4) | 1.03 | 17 (10.8) | 29 (16.1) | 1.03 |
| Allergic rhinitis | 29 (18.4) | 45 (25.0) | 2.17 | 30 (19.4) | 48 (26.7) | 2.49 |
| Food Allergies | 13 (8.2) | 27 (15.0) | 3.70 | 15 (9.7) | 32 (17.8) | 4.53 |
| Postural orthostatic tachycardia syndrome | 1 (0.6) | 5 (2.8) | <sup>F</sup>0.22 | 1 (0.6) | 20 (11.2) | 15.96** |
| Other dysautonomia | 0 (0.0) | 3 (1.7) | <sup>F</sup>0.25 | 0 (0.0) | 13 (7.2) | 11.87** |
| Migraine headaches | 18 (11.4) | 46 (25.6) | 11.00** | 19 (12.2) | 56 (31.1) | 17.28** |
| Mast cell activation syndrome | 0 (0.0) | 1 (0.6) | <sup>F</sup>1.00 | 0 (0.0) | 6 (3.3) | <sup>F</sup>0.03* |
| Myalgic encephalomyelitis/chronic fatigue syndrome | 2 (1.3) | 1 (0.6) | <sup>F</sup>0.60 | 3 (1.3) | 23 (27.8) | 13.68** |
| Fibromyalgia | 0 (0.0) | 8 (4.4) | <sup>F</sup>0.008** | 0 (0.0) | 12 (6.7) | 10.72** |
| Major depression<sup>%</sup> | 23 (14.6) | 23 (12.8) | 0.27 | <sup>+</sup>24 (15.3) | <sup>+</sup>60 (33.5) | 14.83** |
| Bipolar I | 4 (2.5) | 2 (1.1) | <sup>F</sup>0.42 | 4 (2.5) | 2 (1.1) | <sup>F</sup>0.42 |
| Bipolar II | 5 (3.2) | 0 (0.0) | <sup>F</sup>0.02* | 5 (3.2) | 0 (0.0) | <sup>F</sup>0.02* |
| Generalized anxiety disorder<sup>&</sup> | 32 (20.3) | 37 (20.6) | 0.01 | 43 (27.2) | 65 (36.3) | 3.05 |
| Social phobia | 3 (2.7) | 5 (3.1) | <sup>F</sup>1.00 | 3 (2.7) | 5 (3.1) | <sup>F</sup>1.00 |
| Post-traumatic stress disorder<sup>□</sup> | 10 (6.3) | 17 (9.4) | 1.11 | 23 (14.7) | 45 (25.6) | 5.95* |
| Obsessive compulsive disorder | 4 (2.5) | 2 (1.1) | <sup>F</sup>0.42 | 4 (2.5) | 2 (1.1) | <sup>F</sup>0.42 |
| Anorexia nervosa | 3 (1.9) | 2 (1.1) | <sup>F</sup>0.67 | 3 (1.9) | 2 (1.1) | <sup>F</sup>0.67 |
| Bulimia nervosa | 3 (1.9) | 1 (0.1) | <sup>F</sup>0.34 | 3 (1.9) | 1 (0.1) | <sup>F</sup>0.34 |
| Binge eating disorder | 3 (1.9) | 3 (3.7) | <sup>F</sup>0.42 | 3 (1.9) | 3 (3.7) | <sup>F</sup>0.42 |
| Attention-deficit hyperactivity disorder | 4 (3.5) | 5 (3.1) | <sup>F</sup>1.00 | 4 (3.5) | 5 (3.1) | <sup>F</sup>1.00 |
| Current Severity – *M (SD)* |  |  |  |  |  | **F** |
| Depression<sup>%</sup> |  |  |  | 53.11 (10.68) | 59.99 (9.76) | 15.72** |
| Anxiety |  |  |  | 1.80 (1.89) | 2.15 (1.96) | 25.70** |
| Post-traumatic stress disorder |  |  |  | 14.69 (16.15) | 22.93 (17.59) | 5.30* |

<sup>F</sup>Fisher's Exact Test
<sup>%</sup>*Patient Health Questionnaire-9 (PHQ-9)*<sup>23</sup> or *Center for Epidemiology Studies Depression Scale – Revised (CESD-R)*<sup>24</sup> scores converted to Promis Depression T-score equivalents for comparison<sup>25</sup>

### Acute Illness

Participants predominantly experienced mild (40.7%) or moderate (32.6%) acute illness severity, although 5% reported severe illness and developed pneumonia. Compared to the recovered group, long COVID participants experienced greater illness severity and had higher rates of hospitalization (Table 1). During acute illness they more frequently experienced burning sensation, brain fog, chest pain, phantosmia, chanteuse, dyspnea, breathlessness, hypoxia, nausea, diarrhea, constipation, lymphadenopathy, rash, fatigue, headache, myalgias, and chills than those who did not develop long COVID (Figure 1).

**Figure 1.**
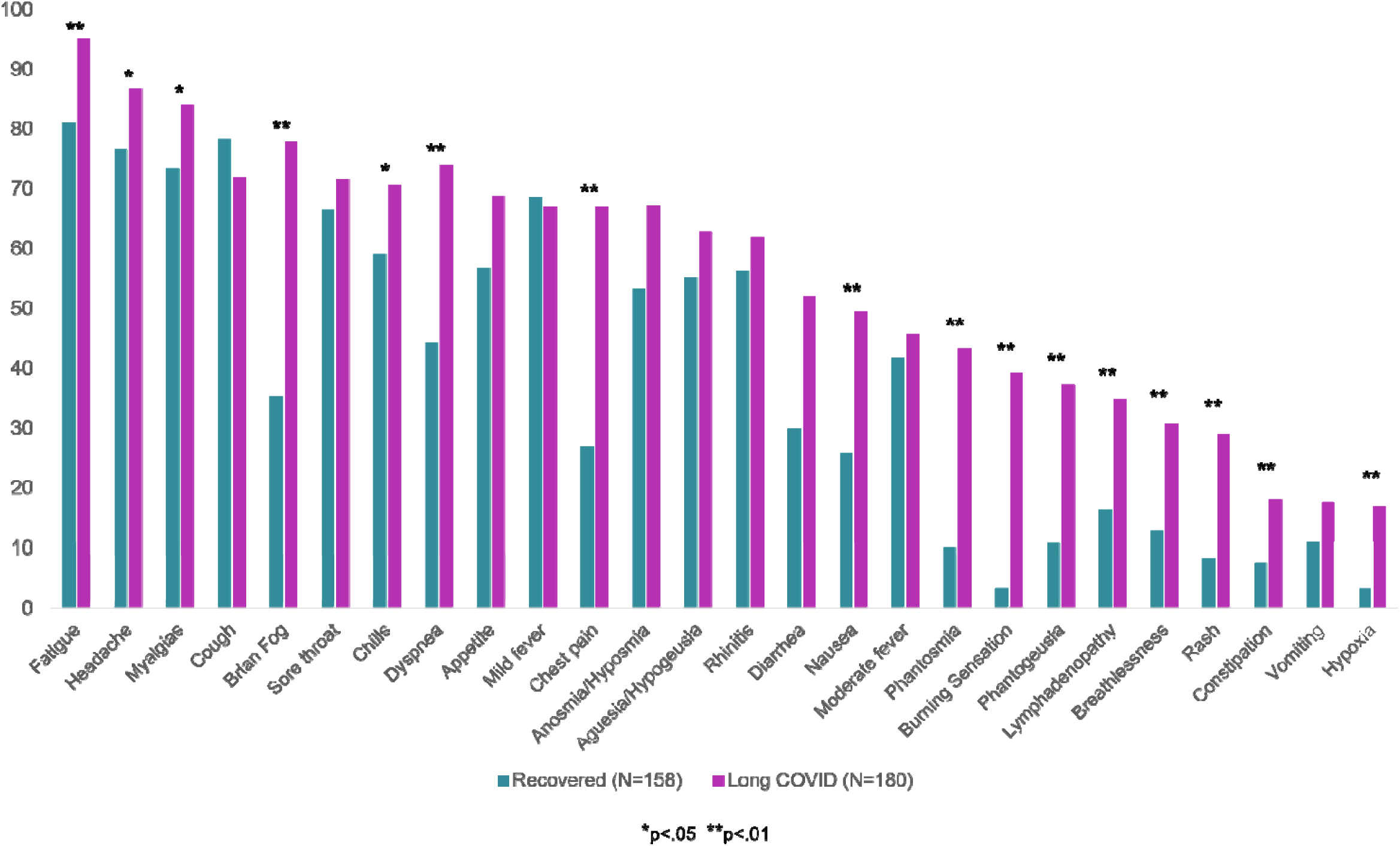
Rates of acute COVID-19 symptoms experienced by recovered and long COVID participants. Displayed are percentage of recovered (green) and long COVID (pink) participants who experienced each symptom during acute COVID-19.

### Post COVID Outcomes

Although new onset health conditions following acute COVID-19 were low across both groups (see Table 2 and supplemental Table 1), those with long COVID reported significantly more myalgia encephalomyelitis/chronic fatigue syndrome (ME/CFS), postural orthostatic tachycardia syndrome, and other dysautonomia diagnoses. As displayed in Figure 2, over 60% of the long COVID group endorsed current fatigue, exercise intolerance, myalgias, headaches, brain fog, and shortness of breath, and at least 40% reported cough, chest pain, chemosensory impairment, and gastrointestinal symptoms.

**Figure 2.**
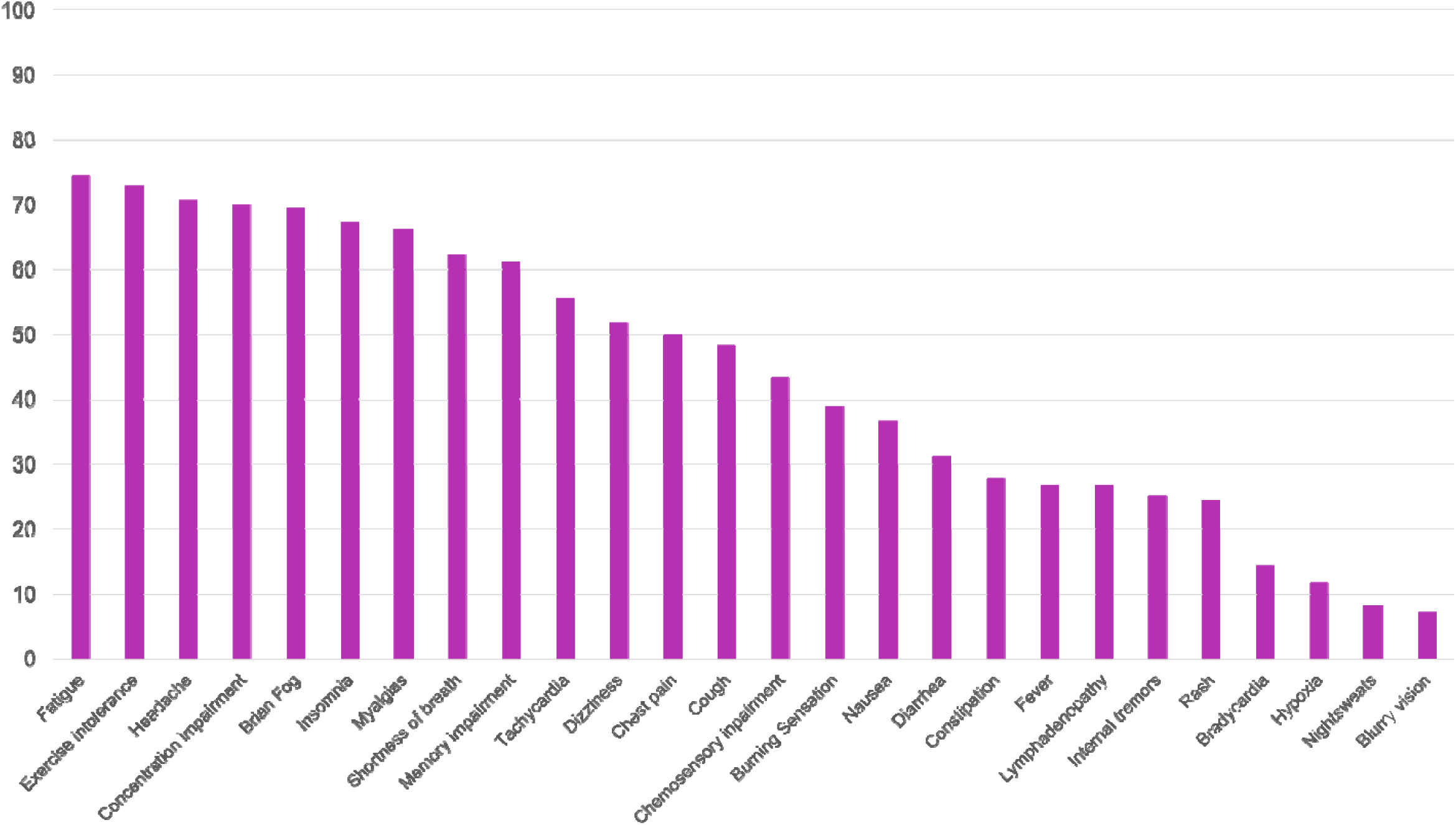
Rates of symptoms experienced in the past month by long COVID participants. Displayed are the percentage of long COVID participants (N=180) who experienced each symptom within the past month.

Although the groups had similar premorbid rates of PTSD, depression, and generalized anxiety, the long COVID group had significantly more post-COVID PTSD and depression (Table 2). Long COVID participants (*F*(1,327)=15.72, *p*<.001, partial η^2^=.046) and males (*F*(1,327)=4.65, *p*=.032, partial η^2^=.014) had higher PTSD severity, with no interaction effects (p=.699). Depression (*F*(1,258)=25.70, *p*<.001, partial η^2^=.073) and generalized anxiety (*F*(1,331)=5.30, *p*=.022, partial η^2^=.016) severity were also higher in the long COVID group, with no sex differences (p’s >.376) or interaction effects (all p’s >.253). Depression (R_s_=.217, p<.001) and PTSD (r_est_=.231, p<.001) were positively correlated with acute illness severity.

### Trauma Exposure

Childhood trauma burden (*F*(1,87)=6.26, *p*=.013, partial η^2^=.038) was significantly higher in the long COVID group, with no sex (p=.940) or interaction (p=.969) effects (Figure 3). The likelihood of developing long COVID increased 3-fold for participants who reported one or more childhood traumatic experiences (OR=3.11, CI, 1.49 to 6.48), while those who endorsed two or more events were 5.6 times more likely to develop the syndrome (OR=5.67, CI, 2.44 to 13.13). Childhood trauma burden was positively correlated with number of premorbid risk factors (r=.298, p<.001) and acute illness severity (r_ds._=.348, p<.001).

**Figure 3.**
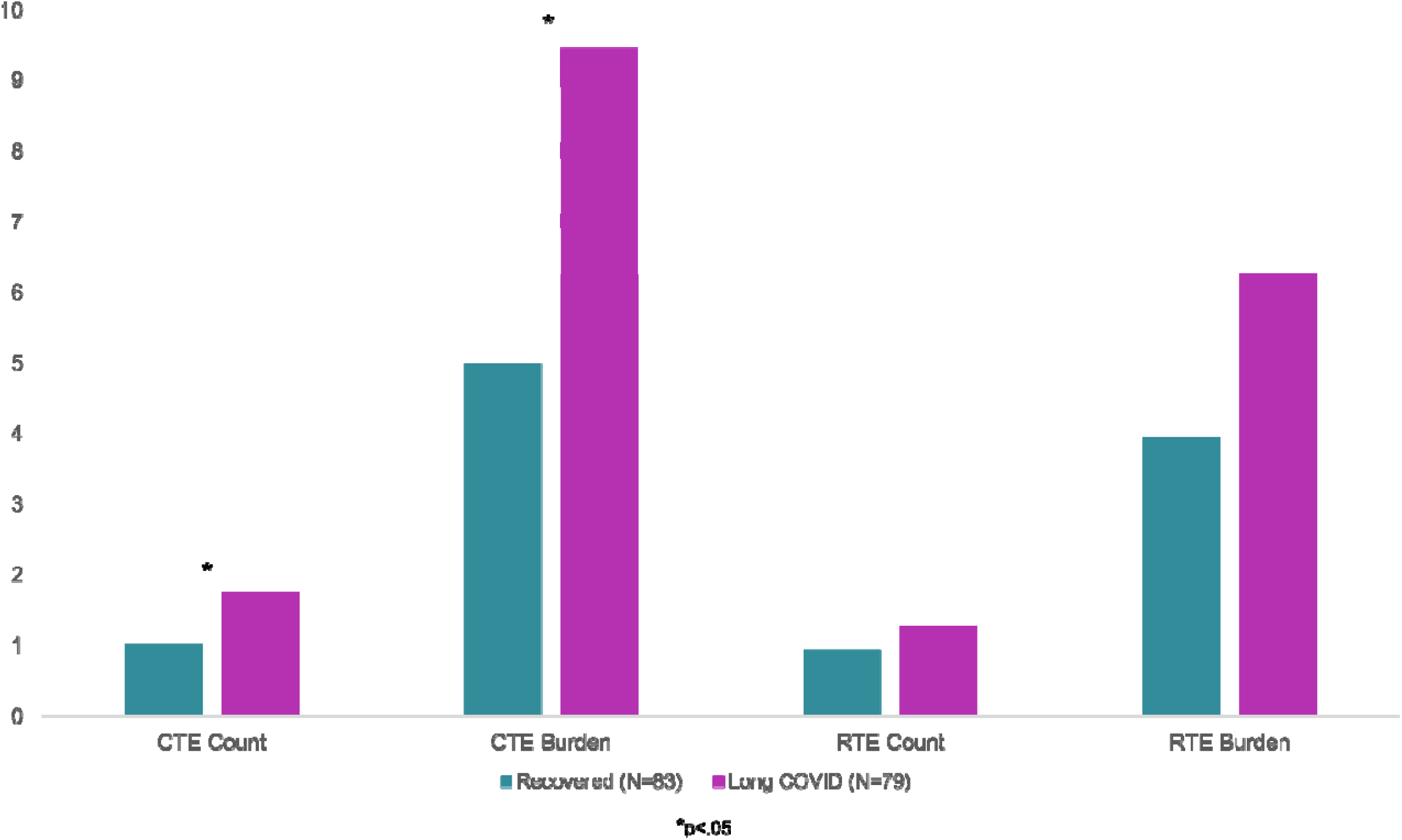
Mean childhood and recent traumatic event count and burden. Displayed are mean childhood traumatic event (CTE) and recent traumatic event (RTE) count and burden for the recovered (green) and long COVID (pink) groups.

Those who experienced one or more traumatic event in the three years prior to contracting COVID-19 were not more likely to develop long-COVID (OR=1.50, CI, 0.78 to 2.90) nor was risk increased for those who reported two or more recent traumatic events (OR=1.88, CI, 0.86 to 4.14). Recent trauma burden was not associated with number of premorbid risk factors or acute illness severity (p’s >.147).

### Predictors of Long COVID

The first logistic regression model demonstrated chest pain (OR=3.35, CI, 1.68 to 6.71), brain fog (OR=2.49, CI, 1.26 to 5.00), phantosmia (OR=3.74, CI, 1.64 to 6.58), and burning sensation (OR=6.69, CI, 2.01 to 22.28) during acute illness, along with female sex (OR=2.54, CI, 1.258 to 5.16) and older age (OR=1.08, CI, 1.05 to 1.11) were independent predictors of long COVID.

The findings were similar in the subset of participants who completed the trauma measures; however, when childhood trauma exposure was entered on step two of the model, only chest pain (OR=8.77, CI, 2.80 to 27.43), brain fog (OR=3.33, CI, 1.16 to 9.57), phantosmia (OR=5.90, CI, 1.40 to 24.86), older age (OR=1.11, CI, 1.06 to 1.16), female sex (OR=4.02, CI, 1.34 to 12.12), and childhood trauma exposure (OR=5.32, CI, 1.44 to 19.68) predicted the syndrome. Together, these variables accurately predicted COVID-19 recovery status for 87% of participants.

## DISCUSSION

Consistent with previous studies, more than half of our sample did not fully recover from COVID-19 within three months, the majority of whom were female. Prolonged immune response is proposed as an underlying mechanism of long COVID,^16^ possibly because females, who are at higher risk for long COVID, produce more robust innate and immune responses than males.^29^ This may prove advantageous against COVID-19 mortality, but be a double-edged sword if such immune responses increase the vulnerability for long COVID.^30^ A recent study found proinflammatory cytokine levels were similar in those who developed long COVID and those who reported full symptom remission four months after SARS-CoV-2 infection, but the long COVID group had higher interferon beta (IFN-β) and interferon lambda (IFN-λ1) levels eight months post-infection.^31^ Further, recovered participants showed naive B and T cells three and eight months post-infection but they were absent at both time-points in the long COVID group, suggesting chronic B and T cell activation.

As hypothesized, we found that exposure to one traumatic event prior to age 17 was associated with a 3-fold increase in long COVID, while those who experienced two or more such events were nearly six times more likely to develop the syndrome. Exposure to childhood and adolescent trauma is known to result in a well-established cascade of maladaptive stress responses primarily mediated by hormonal dysregulation of the hypothalamic-pituitary-adrenal (HPA) axis [reviewed in^32^]. These changes may predispose individuals to altered subjective stress responses as well as certain biological abnormalities such as cortisol elevations, central nervous system hyperresponsiveness, and altered immune response.^19,33^

Myalgia encephalomyelitis/chronic fatigue syndrome (ME/CFS) may be an important illness analogue for understanding the connections between trauma and long COVID, given that the core symptoms of ME/CFS overlap significantly with some features of long COVID. Although predominantly marked by new onset chronic debilitating fatigue, ME/CFS is also characterized by impaired memory and concentration, post-exertional malaise, various hypersensitivities to noxious stimuli, and diffuse pain/headaches without any known medical or psychological cause.^34^ Studies have revealed a range of both hormonal and neurological abnormalities in ME/CFS patients [reviewed in^35^] closely paralleling those aforementioned ones stemming from childhood and adolescent trauma. It should be noted that numerous studies suggest the rates of early life trauma in CFS may exceed 50%.^36-38^ One study showed that greater early childhood trauma exposure was positively correlated with increasing levels of core symptoms in ME/CFS, suggesting a proportional relationship and continuum of severity.^39^

Severe HPA axis dysregulation can cause chronic brain and spinal cord changes resulting in a state of neural vulnerability known as central sensitization.^40^ Characterized by pain hypersensitivity, central sensitization offers a template of chronic risk which interacts poorly with new insults, including, but not limited to, acute illness (such as COVID-19 infection and its associated inflammatory reactions) and psychosocial stressors (such as the known societal and other challenges associated with the pandemic). It should be noted that a large metanalysis comparing various conditions associated with central sensitization found the strongest association between early trauma and ME/CFS,^41^ suggesting a phenotypic propensity for early trauma to inform chronic fatigue and other related symptoms. Consistent with previous reports, myalgias, headaches, chest pain, burning sensation, and internal tremors were frequently endorsed long COVID symptoms in our sample, implicating central sensitization in the pathophysiology of the syndrome. Over half of long COVID participants also developed autonomic dysfunction (e.g., tachycardia, hypertension, orthostatic intolerance, fever, and dyspnea without oxygen desaturation) implicating peripheral dysfunction in long COVID pathophysiology as well.

### Limitations

Our finding that childhood trauma exposure increases long COVID risk is novel, but there are several limitations of this study that must be considered. Participants were recruited online, and it is possible that those who developed long COVID are more invested in participating, potentially inflating the number of individuals with the syndrome, although our rates of long COVID are notably consistent with other reports.^6-10^ We also did not have COVID-19 positivity confirmed for all participants; however, we excluded participants who did not report a positive PCR-test, a clinician diagnosis, or smell/taste loss (which is the strongest indicator of COVID-19 positivity),^21^ minimizing the likelihood of including participants with no SARS-CoV-2 exposure. The cross-sectional design also limits our ability to determine causality, as is the case in any research examining the impact of childhood trauma in adults.

### Conclusion

In sum, our findings provide further evidence that long COVID is not infrequent and that it disproportionately affects females. Childhood trauma exposure may increase long COVID risk via altered immune responses, central sensitization, and disrupted peripheral nervous system function. The widespread prevalence of long COVID may shed light on the shared pathophysiology with other phenotypically similar syndromes that are more rarely diagnosed. Childhood trauma, a crucial social determinant of health, should be routinely assessed in COVID-19 survivors and may be particularly useful for determining prognosis.

## Data Availability

All data produced in the present study are available upon reasonable request to the authors.

