## Supplemental Table 1 for "Childhood Trauma Exposure Increases Long COVID Risk"

**Supplemental Materials**

**Table 1. Additional medical diagnoses**

|  | **Premorbid** | | | **Post-COVID** | | |
| --- | --- | --- | --- | --- | --- | --- |
|  | **Recovered**  **(N=158)** | **Long COVID**  **(N=180)** | **Χ^2^** | **Recovered**  **(N=158)** | **Long COVID**  **(N=180)** | **Χ^2^** |
| Diagnosis – *n (%)* |  |  |  |  |  |  |
| Sleep apnea | 1 (0.6) | 2 (1.1) | **^F^**1.00 | 1 (0.6) | 2 (1.1) | ^F^1.00 |
| Chronic obstructive pulmonary disease | 1 (0.6) | 4 (2.2) | **^F^**0.38 | 1 (0.6) | 6 (3.3) | ^F^0.13 |
| Other pulmonary condition | 2 (1.3) | 2 (1.1) | **^F^**1.00 | 2 (1.3) | 2 (1.1) | ^F^1.00 |
| Congenital heart disease | 2 (1.3) | 1 (0.6) | **^F^**0.60 | 2 (1.3) | 1 (0.6) | ^F^0.60 |
| Coronary artery disease | 1 (0.6) | 2 (1.1) | **^F^**1.00 | 1 (0.6) | 5 (1.1) | ^F^0.22 |
| Congestive heart failure | 0 (0.0) | 2 (1.1) | **^F^**0.50 | 0 (0.0) | 2 (1.1) | ^F^0.50 |
| Pericarditis | 0 (0.0) | 0 (0.0) | **^F^**1.00 | 0 (0.0) | 3 (2.0) | ^F^0.29 |
| Liver Disease | 1 (0.6) | 2 (1.1) | **^F^**1.00 | 1 (0.6) | 3 (1.7) | ^F^0.63 |
| Type I diabetes | 0 (0.0) | 1 (0.6) | **^F^**1.00 | 0 (0.0) | 2 (1.1) | ^F^0.50 |
| Type II diabetes | 2 (1.3) | 7 (3.9) | **^F^**0.18 | 3 (1.3) | 9 (3.9) | 2.27 |
| Anemia | 2 (1.3) | 6 (3.3) | **^F^**0.29 | 2 (1.3) | 6 (3.3) | ^F^0.30 |
| Cancer | 1 (0.6) | 7 (3.9) | **^F^**0.07 | 1 (0.6) | 7 (3.9) | 0.07 |
| Polycystic ovarian syndrome | 1 (0.6) | 2 (1.1) | **^F^**1.00 | 1 (0.6) | 2 (1.1) | ^F^1.00 |
| Colitis | 5 (3.2) | 4 (2.2) | **^F^**0.74 | 5 (3.2) | 7 (3.9) | ^F^0.11 |
| Crohn’s disease | 2 (1.3) | 1 (0.6) | **^F^**0.60 | 2 (1.3) | 1 (0.6) | ^F^0.60 |
| Lupus | 2 (1.3) | 2 (1.1) | **^F^**1.00 | 2 (1.3) | 2 (1.1) | ^F^1.00 |
| Other primary immune deficiency | 3 (1.9) | 2 (1.1) | **^F^**0.67 | 3 (1.9) | 7 (3.9) | ^F^0.35 |
| Celiac disease | 1 (0.6) | 1 (0.6) | **^F^**1.00 | 1 (0.6) | 1 (0.6) | ^F^1.00 |
| Rheumatoid arthritis | 1 (0.6) | 5 (2.8) | **^F^**0.22 | 1 (0.6) | 5 (2.8) | ^F^0.22 |
| Raynaud’s syndrome | 3 (1.9) | 3 (1.7) | 0.03 | 3 (1.9) | 8 (4.4) | 1.65 |
| Guillain-Barré syndrome | 0 (0.0) | 2 (1.1) | **^F^**0.28 | 0 (0.0) | 2 (1.1) | ^F^0.28 |
| Shingles | 2 (1.3) | 7 (3.9) | **^F^**0.18 | 2 (1.3) | 8 (4.4) | ^F^0.11 |
| Recurrent warts/viral skin infections | 4 (2.5) | 3 (1.7) | **^F^**0.71 | 4 (2.5) | 3 (1.7) | ^F^0.71 |
| Multiple sclerosis | 2 (1.3) | 1 (0.6) | **^F^**0.60 | 2 (1.3) | 1 (0.6) | ^F^0.60 |
| Spinal injury | 0 (0.0) | 2 (1.1) | **^F^**0.50 | 0 (0.0) | 2 (1.1) | ^F^0.50 |
| Seizure disorder | 1 (0.6) | 2 (1.1) | **^F^**1.00 | 1 (0.6) | 2 (1.1) | ^F^1.00 |

^F^Fisher’s Exact Test
